# Reduced asymmetry of the hand knob area and decreased sensorimotor u-fiber connectivity in middle-aged adults with autism

**DOI:** 10.1101/2021.07.16.21260584

**Authors:** Janice Hau, Ashley Baker, Chantal Chaaban, Jiwandeep S. Kohli, R. Joanne Jao Keehn, Annika C. Linke, Lisa E. Mash, Molly Wilkinson, Mikaela K. Kinnear, Ralph-Axel Müller, Ruth A. Carper

## Abstract

Individuals with autism spectrum disorder (ASD) frequently present with impairments in motor skills (e.g., limb coordination, handwriting and balance), which are observed across the lifespan but remain largely untreated. Many adults with ASD may thus experience adverse motor outcomes in aging, when physical decline naturally occurs. The ‘hand knob’ of the sensorimotor cortex is an area that is critical for motor control of the fingers and hands. However, this region has received little attention in ASD research, especially in adults after midlife. The hand knob area of the precentral (PrC_hand_) and postcentral (PoC_hand_) gyri was semi-manually delineated in 49 right-handed adults (25 ASD, 24 typical comparison [TC] participants, aged 41-70 years). Using multimodal (T1-weighted, diffusion-weighted, and resting-state functional) MRI, we examined the morphology, ipsilateral connectivity and laterality of these regions. Correlations between hand knob measures with motor skills and autism symptoms, and between structural and functional connectivity measures were also investigated. The right PrC_hand_ volume was greater, and typical leftward laterality of PrC_hand_ and PoC_hand_ volume was lower in the ASD than the TC group. Furthermore, we observed increased mean diffusivity of the right PoC-PrC_hand_ u-fibers in the ASD group. In the ASD group, right PoC-PrC_hand_ u-fiber volume was negatively associated with current autism severity, and positively associated with right PoC-PrC_hand_ functional connectivity (FC). Correlations of hand knob measures were observed with manual dexterity and coordination skills but did not survive multiple comparisons correction. Our findings suggest decreased morphological laterality and u-fiber connectivity of the sensorimotor network involved in hand function in middle-aged adults with ASD. The altered morphology may relate to atypical functional asymmetries found in ASD earlier in life, but additionally, could reflect an overreliance on right hemisphere motor circuits over time. The right PoC-PrC_hand_ u-fibers may underlie compensatory self-regulation of unwanted core motor behaviors seen in ASD.

## Introduction

Autism spectrum disorder (ASD) is characterized by deficits in social communication, atypical patterns of restricted and repetitive behaviors and sensory abnormalities (American Psychiatric Association, 2013). Impairments in the motor domain are not included in the diagnostic criteria (American Psychiatric Association, 2013). The proportion of individuals with ASD at risk of a motor impairment, however, is estimated to be as high as 86.9% (Bhat, 2020; Licari et al., 2020). Motor difficulties in ASD are also seldomly reported by diagnosing clinicians (Licari et al., 2020) and remain largely untreated (Bhat, 2020). Motor impairments in individuals with ASD are observed across a wide range of domains (e.g., gross and fine motor coordination, postural stability, balance and grip strength), and are often related to difficulties in motor planning and increased neural noise in the sensorimotor system (Gowen & Hamilton, 2013). Fine motor impairment in ASD, in particular, is well-documented and has been reported in infancy as early as 6 months (Iverson et al., 2019; Libertus et al., 2014; Sacrey et al., 2018), at the toddler stage (Landa & Garrett-Mayer, 2006; Lloyd et al., 2013), in childhood (Fuentes et al., 2009; Green et al., 2009; Jasmin et al., 2009; Provost et al., 2007), adolescence (Fuentes et al., 2010; Pan, 2014) and into middle-age (Linke et al., 2019; Thompson et al., 2017; Travers et al., 2015, 2017).

Motor skills are essential for daily living activities required to independently care for oneself, such as eating, dressing, and walking. In children and adults with ASD, positive correlations of manual motor skills with daily living skills, and of motor coordination with daily living skills and adaptive behavior, have been reported (Bremer & Cairney, 2018; Travers et al., 2017). In aging, the ability to perform these activities is widely used as an indicator of health and disability outcomes (Katz, 1983). Manual motor skills are also found to be robust predictors of functional disability (Rantanen et al., 1999), dependency (Ostwald et al., 1989) and even cognitive impairment (Curreri et al., 2018) in the elderly. In individuals with ASD, there is some evidence that manual motor skills may lag increasingly behind with age (Travers et al., 2017). In an accelerated longitudinal study involving individuals with ASD aged 5-39 years at their first visit, Travers and colleagues (2017) noted that after around age 15 years, grip strength and finger tapping speed in the ASD group diverged further from a typically developing group (with a larger gap in performance than at a younger age). Thus, early motor impairments may be compounded by increased motor problems in adolescence and adulthood in ASD. Reports of an increased prevalence of movement disorders (e.g., parkinsonism) in middle-aged and older adults with ASD have emerged (Croen et al., 2015; Starkstein, Gellar, Parlier, Payne, & Piven, 2015); however, the literature on aging in ASD remains extremely limited (Piven & Rabins, 2011).

The anatomical ‘hand knob’ landmark is a characteristic knob-like shape formed by the central sulcus that defines subareas of the pre and postcentral gyri (Yousry et al., 1997). The hand knob areas elicit hand movements when electrically stimulated (Penfield & Boldrey, 1937) and consistently show activation during hand movements (Sun et al., 2016; Yousry et al., 1997) in the contralateral hemisphere. Although they are considered to be primarily specialized for hand motor execution, there is evidence of involvement in premotor functions beyond basic motor execution, such as motor planning and coordination. For example, cytoarchitecture of the dorsal premotor cortex (corresponding to Brodmann area 6) extends to portions of the rostral precentral gyrus including the hand knob area (Geyer et al., 1996; Geyer, Matelli, Luppino, & Zilles, 2000; Rademacher et al., 2001; Rademacher, Rademacher, Caviness, Steinmetz, & Galaburda, 1993; L. E. White et al., 1997). Stimulation studies of the pre and postcentral hand knob areas show substantial overlap between representations of the hand and other body parts (e.g., mouth, arm, face) in typical individuals (Catani, 2017; Penfield & Boldrey, 1937), and a recent study proposed that the hand knob area may be involved in compositional coding of movement across limbs (Willett et al., 2020).

Neuroimaging studies on the hand knob area have mainly been limited to the topics of manual preference (Amunts et al., 1996; Dassonville et al., 1997; S Rose et al., 2012; Volkmann et al., 1998), hand paresis (Gass, Szabo, Behrens, Rossmanith, & Hennerici, 2001; Peters et al., 2009; Stephen Rose, Guzzetta, Pannek, & Boyd, 2011) and neurophysiology (Hallett, 2007). To our knowledge, only one MRI study has specifically investigated the hand knob area in ASD, focusing exclusively on the short-range association white matter tracts (i.e., u-fibers) (Thompson et al., 2017). Using deterministic tractography, Thompson and colleagues found evidence of compromised microstructure, specifically reduced fractional anisotropy and increased mean and radial diffusivity, bilaterally in the u-fibers connecting ipsilateral postcentral and precentral hand knob area in right-handed adults (aged 18-45 years) with ASD relative to a control group. Of the limited literature on middle-aged adults with ASD, reduced resting-state functional connectivity of intra and interhemispheric primary sensorimotor and premotor cortical areas has been observed by our group (Linke et al., 2019). This appears to be consistent with the reduced postcentral to precentral u-fiber structural integrity reported in the slightly younger adults with ASD. However, whether these differences in the u-fiber microstructural indices in ASD continue to be seen in middle-age, and what the relationship is between structural and functional connectivity measures of these regions, is unknown.

Although not specifically examining the hand knob area, previous MRI studies have found grey matter abnormalities in the pre and postcentral gyri in ASD. Meta-analyses of voxel-based morphometry studies involving a wide age range from children to adults (including middle-age) have reported decreased grey matter density of the left precentral gyrus (Cauda et al., 2011; Nickl-Jockschat et al., 2012) and left postcentral gyrus (DeRamus & Kana, 2015) in ASD compared with typical development, and increased grey matter density of the right precentral gyrus in adults (18-52 years) with ASD (DeRamus & Kana, 2015). Moreover, an altered trajectory with age in grey matter density of right precentral gyrus is observed in ASD, declining linearly from age 8 to 50 years while showing a quadratic curve in a typically developing group, with atypically increased grey matter density in ASD in early and middle adulthood (Greimel et al., 2013). This pattern of results raises the possibility of reduced grey matter asymmetry in the pre- and postcentral gyri in adults with ASD. Previous studies on morphological asymmetry, however, have not revealed any group differences in asymmetry in these regions in ASD (Floris, Lai, et al., 2016; Postema et al., 2019). It is possible, however, that laterality varies in different portions of the relatively large regions of the pre and postcentral gyri, and the effect of reduced asymmetry is more localized (e.g., within the hand knob).

The present multimodal MRI study is the first to examine the morphology, structural and functional connectivity, and asymmetry of the hand knob area and links with both motor skills and autism symptoms in middle-aged adults with ASD. Based on the previous literature, we hypothesized reduced asymmetry in volume (accompanied by decreased left and/or increased right volume) in the pre-(PrC_hand_) and postcentral (PoC_hand_) hand knob area, and decreased pre- to post-central hand knob (PoC-PrC_hand_) connectivity in ASD. We also hypothesized that greater leftward laterality of structure (i.e., hand knob volume and u-fiber tract integrity) and functional connectivity would be correlated with improved motor skills and less severe autism symptoms. Additionally, we examined the superior and inferior segments of the pre- and postcentral gyri above and below the hand knob area, in order to determine the specificity of any diagnostic group differences within these gyri. Links between PoC-PrC_hand_ structural and functional connectivity were also investigated.

## Methods

### Participants

Seventy-three participants (35 ASD, 37 typical comparison [TC]), aged 40-70 years, were recruited from San Diego County communities through referrals from autism clinics, service providers (e.g., day programs, group homes) and local advertisement as part of an ongoing longitudinal study. Only individuals with idiopathic ASD and without epilepsy were eligible for participation. TC participants had no personal or family history of autism. All participants had no personal history of other genetic or neurological conditions or serious mental illness. Seventeen datasets were excluded after quality assessment of the imaging data (12 were affected by a hardware issue, 2 scans were incomplete, 3 had low quality anatomical images due to noise, motion or artifacts – see below) and 3 participants were excluded for incidental brain findings. As our study included laterality measures for motor regions, left-handed participants (1 ASD, 2 TC) were excluded. The final sample consisted of 25 ASD and 24 TC participants. For the fMRI analyses, 7 participants were further excluded (4 ASD and 2 TC participants for image artifacts or excessive head motion; 1 ASD participant had no fMRI data). Behavioral measures, including BMAT scores, for the majority of these participants were previously presented in (Linke et al., 2019).

All ASD diagnoses were confirmed by a clinical psychologist according to DSM-5 criteria (American Psychiatric Association, 2013), and supported by module 4 of the Autism Diagnostic Observation Schedule, 2^nd^ edition (ADOS-2) (Lord et al., 2002) (current ADOS scores were not available for one participant due to administrator error, however this individual met ASD criteria on a research-related ADOS within the preceding 5 years). Intelligence quotient (IQ) was assessed with the Wechsler Abbreviated Scale of Intelligence, 2^nd^ edition (WASI-II) (Wechsler, 2011). Dichotomous hand preference (left/ambidextrous or right) was determined for all participants based on a personal history questionnaire completed by the participant or caregiver in response to the question “With which hand does s/he eat, brush teeth, write, etc.?”, in line with (Perelle & Ehrman, 2005). The Edinburgh Handedness Inventory (EHI) (Oldfield, 1971) was used to measure degree of handedness and was available for 23 ASD and 23 TC participants. The EHI consists of a self-report checklist in which participants indicate their hand preference (left, right, or if really indifferent, both left and right) across ten different actions (e.g., “write a letter with a pencil”, “throw a baseball”, “strike a match”), and a final score is calculated as follows: (# right - # left) / (# right + # left) x 100. The self-reported EHI scores corresponded to dichotomous hand preference for all participants who completed the EHI. Motor function was assessed using the short form version of the Bruininks Motor Ability Test (BMAT) (Bruininks & Bruininks, 2012). Five subcategories of motor behavior were evaluated: fine motor (e.g., drawing a line through a curved path), manual dexterity (e.g., transferring pennies), coordination (e.g., bouncing and catching a ball), balance and mobility (e.g., standing on one leg), and strength and flexibility (e.g., grip strength).

The study was approved by the institutional review boards of the San Diego State University and University of California, San Diego. All participants were screened for MRI contraindications (e.g., claustrophobia, ferrous material in body). All participants or their conservators provided written informed consent before participation, and participants were compensated for their time.

### Image acquisition

MRI data were collected at the Center for Functional Magnetic Resonance Imaging (CFMRI, University of California, San Diego) on a 3T GE Discovery MR750 scanner using a 32-channel head coil. A high resolution anatomical T1-weighted image (repetition time [TR]=8.776 ms, echo time [TE]=3.656 ms, 8° flip angle, 320 x 320 matrix, 0.8mm^3^ resolution) was acquired using an MPRAGE sequence with real-time prospective motion correction (PROMO; White et al., 2010). Diffusion-weighted images were acquired with 46 non-colinear directions at b=1500 (13 ASD and 9 TC participants) or 1000 s/mm^2^ (12 ASD and 15 TC participants) and 6 interspersed volumes at b=0 s/mm^2^ (TR=4000ms, TE=89ms, multi-band factor=3, 27 slices/band, 24cm field of view [FOV], 1.7 mm^3^ resolution) with reversed (anterior-posterior) phase encoding to correct for susceptibility distortions and improve signal-to-noise ratio. Resting-state functional MRI images were acquired using a multi-band EPI sequence (2 runs x 6 minute duration each, TR=800ms, TE=35ms, multi-band factor=8, 52° flip angle, 20.8cm FOV, 2mm^3^ resolution) with a 20s reverse phase-encoded calibration scan pair using identical imaging parameters to correct for susceptibility-induced distortions. Before each functional MRI scan, participants were given the following instructions: “Keep your eyes on the cross. Let your mind wander, relax, but please stay as still as you can. Try not to fall asleep.” Eye status was monitored throughout the duration of the scan using an in-bore camera to ensure participant compliance.

### Data preprocessing

FreeSurfer 5.3.0 was used to preprocess the anatomical images using the standard pipeline (i.e., skull stripping, intensity normalization, gray-white matter segmentation) and to perform cortical surface reconstruction (Dale et al., 1999; Fischl et al., 1999). The anatomical images and surfaces were visually inspected, and any images with inaccurate surfaces or excessive image artifacts (e.g., ghosting, ringing) were excluded. For both diffusion-weighted and functional MRI data, susceptibility-induced off-resonance fields were estimated and corrected from the reversed phase-encoded image pairs using TOPUP (Andersson et al., 2003). For the diffusion-weighted images, we first applied denoising by eliminating the noise component in the patch-level PCA-domain (Veraart et al., 2016). Eddy current and susceptibility-induced distortions, inter-and intra-volume head motion and signal dropout incorporating information from both individual slices and multi-band slice groupings (Andersson et al., 2017) were then corrected in a single interpolation step using EDDY from the FMRIB Software Library (FSL) 5.0.11 (Andersson et al., 2016; Andersson & Sotiropoulos, 2016). The b-vectors were rotated according to the motion realignment. Functional MRI preprocessing included rigid-body realignment for motion correction and non-linear registration to Montreal Neurological Institute (MNI) space (SPM12) and scrubbing of motion outliers, band-pass filtering using a temporal filter of 0.008-0.08 Hz, and nuisance regression (including motion parameters and their derivatives as well as time-series derived from cerebrospinal fluid and white matter) using the Functional Connectivity (Conn) Toolbox (Whitfield-Gabrieli & Nieto-Castanon, 2012).

For purposes of group matching, in-scanner head motion was quantified in two ways for the diffusion-weighted images: average root mean squared displacement excluding in the phase-encoding direction across all brain voxels with respect to the first volume (absolute RMSD) and with respect to the preceding volume (relative RMSD). RMSD was also calculated from the fMRI realignment parameters and averaged across runs to quantify head motion during the functional MRI scans.

### Delineation of the hand knob regions of interest

Given the interindividual variability in the location (Sun et al., 2012) and morphology (Caulo et al., 2007) of the hand knob along the central sulcus, the regions of interest (ROIs) for the pre (PrC_hand_) and postcentral (PoC_hand_) hand knob areas were semi-manually delineated in each participant. Automatic parcellations of the pre and postcentral gyri (Desikan et al., 2006) were performed for each participant and used as starting regions, from which PrC_hand_ and PoC_hand_ were subsequently delineated, to ensure that the final ROIs correspond with consistent pre and post-central cortical boundaries and with grey matter tissue. From the starting parcels, manual segmentations of the hand knob ROIs were performed by one trained operator (AB). Fsleyes (McCarthy, 2018) was used to visualize and edit the ROIs with the participants’ T1-weighted image underlay. First, the hand knob landmark, commonly identified as an omega (but also epsilon or null) shape on the precentral gyrus (Caulo et al., 2007), was located in the axial plane in each hemisphere (Figure 1A). Then the starting regions were manually edited by removing areas surrounding the hand knob (i.e., supero-posterior and infero-anterior to the hand knob [e.g., omega] landmark) until only the hand knob remained (Figure 1B). As the hand knob was most identifiable on the precentral gyrus, delineation of the precentral hand knob (PrC_hand_) was performed first and the boundaries of the postcentral hand knob (PoC_hand_) were extrapolated following a perpendicular line from the superior and inferior limits of PrC_hand_ to the adjacent postcentral gyrus. The T1-weighted images were rigid-body registered to diffusion space using a boundary-based approach (Greve & Fischl, 2009) and the parcellated regions were registered using the same registration matrix with nearest neighbor interpolation. All operators were trained by an experienced anatomist (JH) until agreement was reached on the location and spatial extent of the ROIs in 10 practice participants. All operators were blind to the participants’ diagnostic group.

**Figure 1.**
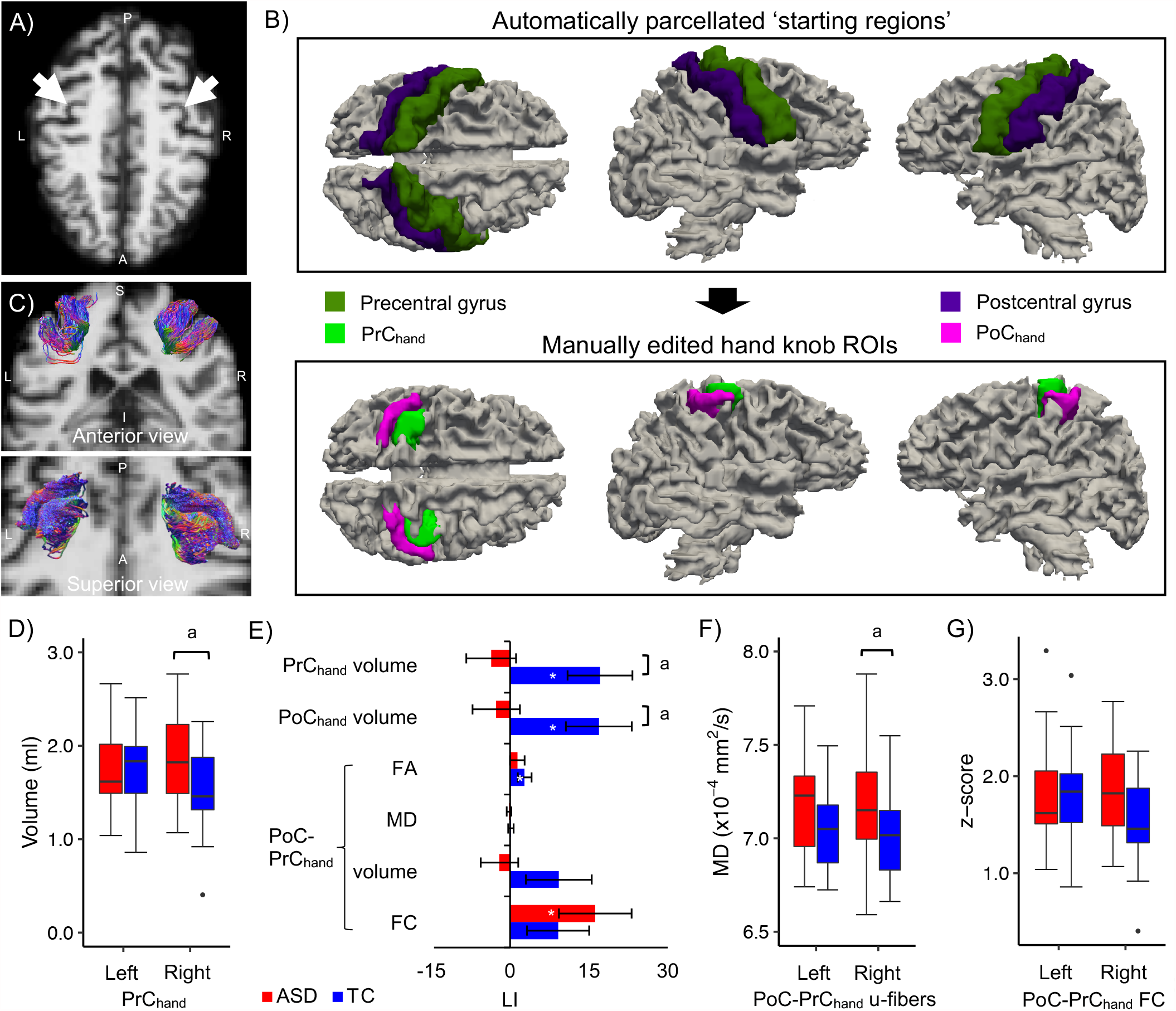
A-C) Identification and delineation of the hand knob regions and u-fibers. A) The typical omega-shaped landmark (white arrows) used to identify the hand knob in the axial plane of a representative participant with ASD (aged 46-50 years). B) The automatically parcellated pre and postcentral regions (upper row) were manually edited to define the pre (PrC_hand_) and postcentral (PoC_hand_) hand knob regions of interest (lower row). C) Streamline tractography of the u-fiber tracts connecting ipsilateral PoC_hand_ and PrC_hand_ from the same participant. D-G) Alterations in hand knob morphometry, laterality and PoC-PrC connectivity in middle-aged adults with ASD. The plots show D) increased right PrC_hand_ volume, E) reduced laterality of PrC_hand_ and PoC_hand_ knob volume (LI of other hand knob measures are also shown), and F) increased MD in the right PoC-PrC_hand_ u-fiber tract in the ASD group. G) Medium size group effects were also observed for left PoC-PrC_hand_ u-fiber MD (increased in ASD) and right PoC-PrC_hand_ FC (decreased in ASD). ^a^ *q*<.10 significance level; ^*^ LI measures that differed from zero (*p*<.05, uncorrected). Standard errors of the means are shown in the bar plot.

To assess intra-rater reliability, the hand knob regions were delineated a second time by author AB in a subset of 15 participants. To assess inter-rater reliability, a second trained operator (CC) delineated hand knob regions in the same subset of participants. Intra-class correlation coefficients (ICC) with two-way random effects showed good overall intra-rater reliability between hand knob volume measurements (PrC_hand_: average measure ICC=.647, p=.003; PoC_hand_: average measure ICC=.823, p<.001) and their laterality indices [LI] (LI PrC_hand_: average measure ICC=.755, p=.006; LI PoC_hand_: average measure ICC=.828, p=.001). Between-rater reliability was also good overall (PrC_hand_: average measure ICC=.667, p=.002; PoC_hand_: average measure ICC=.855, p<.001; LI PrC_hand_: average measure ICC=.770, p=.005.; LI PoC_hand_: average measure ICC=.785, p=.003). Additionally, we calculated overlap coefficients (OC) for PrC_hand_ and PoC_hand_ which showed good spatial correspondence within (PrC_hand_: mean OC [min, max]=.85 [.53, .99]; PoC_hand_: .91 [.70, .99]) and between (PrC_hand_: .87 [.74, .99]; PoC_hand_: .88 [.55, .96]) raters.

Since the hand knob ROIs were delineated in the native diffusion space and therefore could be subject to variations in brain alignment with respect to the cardinal axes, we extracted the root mean squared deviation in roll, pitch and yaw rotations (RMSD_rot_) from the affine transformations between diffusion and MNI space for each participant. Independent t-tests showed that the groups marginally differed in RMSD_rot_ (*p*=.082; TC mean [SD]: 9.66 [3.60]; ASD: 7.97 [3.05]). This measure was included as an additional covariate for the anatomical measures in our statistical model to control for brain image rotation.

### Fiber tractography and diffusion scalar maps

Fiber tractography was performed using the MRtrix3 software package (Tournier et al., 2019). Response functions were estimated for each tissue type (grey matter, white matter and cerebrospinal fluid) using an unsupervised algorithm (Dhollander et al., 2019). Fiber orientation distributions were estimated using multi-tissue constrained spherical deconvolution (Jeurissen et al., 2014), and intensity normalization was applied across tissue-specific distributions to minimize spurious peaks driven by single-tissue intensities. Sensorimotor (PoC-PrC) u-fibers were obtained by performing anatomically constrained probabilistic tractography (Smith et al., 2012) on the fiber orientation distributions using ipsilateral PrC_hand_ and PoC_hand_ as both seed and target regions, that were reversed to track in both directions to control for biases in tracking direction, with 5000 randomly placed seeds per voxel (minimum length=10mm, maximum length=100mm, 45° angle threshold, step size=1mm). The reverse-track pairs were combined to create the final tracts (for an example, see Figure 1C) and were consistent with descriptions of these u-fibers in previous studies (Catani et al., 2012; Guevara et al., 2017; Pron et al., 2021; Rojkova et al., 2015; Viganò et al., 2019).

The diffusion data were fitted to the tensor model at each voxel using weighted least squares to create diffusion tensor imaging scalar maps (fractional anisotropy [FA] and mean [MD],). The scalar maps were harmonized across acquisition protocols (b-value) using an empirical Bayes-based method (Johnson et al., 2007) shown to be robust for small sample sizes and biased samples (ComBat; Fortin et al., 2017).

### Morphological, connectivity and laterality measures

The PrC_hand_ and PoC_hand_ ROI volumes of each participant were calculated as the total volume of voxels occupied by each ROI in the subjects’ diffusion space using FSLstats. In subsequent tests of surface area (SA) and cortical thickness (CT), the regions were re-registered to anatomical space, projected to the cortical mid-surface, smoothed with a Gaussian kernel (full width at half maximum=6mm) and threshold at 20% probability in FreeSurfer. The threshold was chosen to maximize areal coverage while minimizing overlap between PrC_hand_ and PoC_hand_. SA and CT were averaged across the ROI.

The u-fiber tract measures were extracted by averaging the mean value along streamlines from the diffusion scalar maps. The volume of each u-fiber tract was calculated as the total volume of voxels occupied by the tract.

Blood oxygenation level-dependent time-series from the same individual-subject ROIs (warped to MNI standard space) were averaged across voxels, and FC was estimated between ipsilateral PrC_hand_ and PoC_hand_ using Fisher *z*-transformed bivariate Pearson correlations.

To analyze laterality of the anatomical and connectivity features, a laterality index (LI) was calculated for all measures as follows: 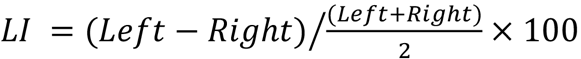, where positive values indicate lateralization in the dominant (left) hemisphere for motor function. Note that this differs from the EHI scores where positive values indicate dominant (right) hand motor function.

Additionally, for each participant, total brain volume was calculated from the tissue maps segmented from the b0 image using FSL’s FAST (Zhang et al., 2001) as the total volume of grey and white matter voxels.

In order to determine the anatomical specificity of our findings, analyses were performed on the pre- and postcentral gyri parcels above (PrC_upper_ and PoC_upper_, respectively) and below (PrC_lower_ and PoC_lower_, respectively) the hand knob using the same methods for extracting and analyzing the hand knob measures. (See Supplementary Figures 1-2 for the upper and lower PrC and PoC parcels and upper and lower PoC-PrC u-fibers in relation to the hand knob parcels in a representative subject).

### Statistical analysis

Independent samples *t*-tests and Chi-square tests were used to compare diagnostic groups on demographic, confounding variables (e.g., in-scanner head motion) and motor performance. One-way analyses of covariance (ANCOVAs) were used to test for effects of diagnosis controlling for age (morphological measures); age, relative RMSD and a dummy variable coding for b-value protocol (u-fiber measures); and age and average RMSD (FC measures). Total brain volume (TBV) (morphological measures) and tract volume (diffusion measures), known to bias diffusion measures due to partial volume effects (Vos et al., 2011), were additionally included in the model to test potential confounding factors. One-sample *t*-tests (two-tailed) against a null-hypothesis of 0 were used to test for laterality of each hand knob measure in the diagnostic groups separately, in addition to analyzing LI for group differences. One TC participant was deemed an outlier (>3 standard deviations from the group mean) and was removed from subsequent analyses of the left PoC-PrC_hand_ u-fiber tract and the tract’s LI. We controlled for multiple comparisons using the false discovery rate (FDR) (Benjamini & Hochberg, 1995) at *q*<.10 across all hand knob measures (18 comparisons).

Pearson partial correlations were performed between the hand knob measures and BMAT subscales (Manual Dexterity, Coordination, and Strength and Flexibility) for each group separately, partialing out the same variables as described for the ANCOVAs above. Due to the limited range of scores in the Fine Motor and Balance and Mobility BMAT subscales in both groups, these were not included in our analyses. Given the ordinal nature of the ADOS-2 measures, Spearman rank-order partial correlations were performed between hand knob measures and the Social Affect, Restricted and Repetitive Behavior [RRB], and Comparison [CS] scores of the ADOS-2 in the ASD group, partialling out the same variables as described for the ANCOVAs above. We applied FDR correction on 18 hand knob measures for each subscale per group.

To explore relationships between structural and functional connectivity, partial Pearson correlations were performed between sensorimotor hand knob u-fiber and functional connectivity measures ipsilaterally, controlling for age, relative RMSD (dMRI motion), average RMSD (fMRI motion) and b-value protocol in each diagnostic group, separately. We applied FDR correction on 3 tract measures [FA, MD and volume] ⨯ 3 FC measures [left, right and LI] (9 comparisons) per diagnostic group.

Statistical analyses were conducted in R (Core R Team, 2019) and plots were created using the package *ggplot2* (Wickham, 2016).

## Results

The diagnostic groups did not differ on age, sex, degree of handedness, IQ, motion dropout or absolute RMSD (Table 1; all *p*s>.05). The ASD group had lower performance in Manual Dexterity, Coordination and Strength and Flexibility compared with the TC group (Table 1). As the groups marginally differed in relative RMSD, this motion measure was included as a covariate in analyses examining diffusion-weighted imaging measures. While the percent difference between groups in motion dropout was 43.1%, it was well below 1% of all slices in all participants, and affected slices were replaced with predicted values during preprocessing with the EDDY tool (Andersson et al., 2016). The groups did not differ in TBV. The sample included in the FC analyses did not differ between groups on age (*p*=.641), sex (*p*=.691), degree of handedness (*p*=.464), IQ (non-verbal: *p*=.752, verbal: *p*=.328) or motion (*p*=.170).

**Table 1.** Participant characteristics and group matching.

| | ASD (n = 25) | | TC (n = 24) | | $p^*$ | % diff |
| --- | --- | --- | --- | --- | --- | --- |
| | Mean ± SD | [range] | Mean ± SD | [range] | | |
| Sex (M/F) | 20/5 |  | 21/3 |  | 0.70 | -- |
| Age (years) | 51.7 ± 6.9 | [41-67] | 51.6 ± 7.0 | [41-70] | 0.95 | 0.1 |
| EHI<sup>†</sup> | 81 ± 30 | [11-100] | 86 ± 18 | [40-100] | 0.47 | -1.6 |
| WASI-2 Verbal IQ | 103 ± 24 | [52-160] | 114 ± 14 | [85-144] | 0.07 | -2.4 |
| Non-verbal IQ | 108 ± 22 | [56-138] | 112 ± 12 | [90-135] | 0.36 | -1.1 |
| Full-scale IQ | 106 ± 23 | [51-143] | 115 ± 12 | [92-138] | 0.09 | -2.0 |
| ADOS-2<sup>‡</sup> Social Affect | 10.2 ± 3.6 | [5-19] | -- | -- | -- | -- |
| RRB<sup>‡</sup> | 3.8 ± 1.8 | [1-8] | -- | -- | -- | -- |
| Comparison Score<sup>‡</sup> | 7.4 ± 1.8 | [3-10] | -- | -- | -- | -- |
| BMAT<sup>§</sup> Fine Motor | 13 ± 1 | [8-15] | 14 ± 1 | [12-14] | 0.11 | -0.9 |
| Manual Dexterity | 12 ± 3 | [2-16] | 14 ± 1 | [12-17] | <.01 | -4.3 |
| Coordination | 9 ± 2 | [4-10] | 10 ± 1 | [7-10] | <.01 | -3.2 |
| Strength & Flexibility | 14 ± 4 | [6-18] | 17 ± 2 | [10-18] | <.01 | -4.9 |
| Balance & Mobility | 9 ± 1 | [7-9] | 9 ± 1 | [6-9] | 0.89 | -0.1 |
| Total brain volume (cm<sup>3</sup>) | 1516 ± 185 | [1085-1783] | 1493 ± 138 | [1096-1686] | 0.61 | 0.4 |
| Motion dropout (% slices) | 0.05 ± 0.13 | [0-0.64] | 0.01 ± 0.12 | [0-0.06] | 0.13 | 43.1 |
| dMRI absolute RMSD | 1.34 ± 1.17 | [0.34-5.34] | 1.13 ± 0.59 | [0.43-2.78] | 0.44 | 4.2 |
| dMRI relative RMSD | 0.43 ± 0.26 | [0.16-1.03] | 0.31 ± 0.11 | [0.17-0.59] | 0.05 | 7.8 |
| fMRI RMSD (average across runs)<sup>¶</sup> | 0.10 ± 0.05 | [0.04-0.19] | 0.09 ± 0.03 | [0.04-0.19] | 0.17 | 4.7 |

\*T-tests; Chi-Square test for sex.
<sup>†</sup>EHI unavailable for 3 participants (2 ASD, 1 TC).
<sup>‡</sup>ADOS-2 scores unavailable for 1 ASD participant due to administrator error (see text).
<sup>§</sup>BMAT was not completed in 2 participants (1 ASD, 1 TC).
<sup>¶</sup>2 participants (1 ASD, 1 TC) were excluded for excessive image artifacts, 4 participants were excluded to match for head motion (3 ASD, 1 TC) and 1 ASD participant did not have functional MRI data.
Abbreviations: EHI=Edinburgh Handedness Inventory; WASI-2=Wechsler Abbreviated Scale of Intelligence, 2<sup>nd</sup> edition; ADOS-2=Autism Diagnostic Observation Scale, 2<sup>nd</sup> edition; RRB=Restricted and Repetitive Behaviors; BMAT=Bruininks Motor Assessment Scale – Short form; RMSD=average voxel-wise root mean square displacement (head motion); diff=difference between groups.

### Hand knob morphology and laterality

The hand knob ROIs were successfully delineated in all participants and the proportion of each morphological variant (80.6% omega, 17.3% epsilon and 2.0% null; Supplementary Table 1) was in line with previous studies(Caulo et al., 2007).

ANCOVAs indicated that right PrC_hand_ volume was significantly increased in the ASD compared with the TC group (Figure 1D; Table 2), while no group difference was found for left PrC_hand_ volume.

**Table 2.** Results of the ANCOVAs performed on the hand knob measures and their group means and standard deviations.

| Region | Measure | df | *p* | *q* | Partial $\eta^2$ | ASD mean ± SD | TC mean ± SD |
| --- | --- | --- | --- | --- | --- | --- | --- |
| Left PrC<sub>hand</sub> | Volume | 1, 46 | 0.962 | 0.97 | <0.001 | 1.80 ± 0.52 | 1.79 ± 0.49 |
| Right PrC<sub>hand</sub> | Volume | 1, 46 | 0.012 | 0.08<sup>a</sup> | 0.129 | 1.85 ± 0.46 | 1.52 ± 0.43 |
| Left PoC<sub>hand</sub> | Volume | 1, 46 | 0.262 | 0.39 | 0.027 | 2.32 ± 0.48 | 2.52 ± 0.70 |
| Right PoC<sub>hand</sub> | Volume | 1, 46 | 0.210 | 0.38 | 0.034 | 2.39 ± 0.55 | 2.17 ± 0.67 |
| PrC<sub>hand</sub> | LI Volume | 1, 46 | 0.011 | 0.08<sup>a</sup> | 0.133 | -3.6 ± 24.7 | 17.1 ± 30.2 |
| PoC<sub>hand</sub> | LI Volume | 1, 46 | 0.015 | 0.08<sup>a</sup> | 0.123 | -2.6 ± 22.5 | 16.9 ± 30.7 |
| Left PoC-PrC<sub>hand</sub> u-fibers | FA | 1, 43 | 0.143 | 0.29 | 0.042 | 0.36 ± 0.02 | 0.37 ± 0.02 |
| | MD | 1, 43 | 0.039 | 0.13 | 0.085 | 7.16 ± 0.24 | 7.03 ± 0.22 |
| | Volume | 1, 43 | 0.972 | 0.97 | <0.001 | 9.94 ± 2.37 | 9.65 ± 1.97 |
| Right PoC-PrC<sub>hand</sub> u-fibers | FA | 1, 44 | 0.899 | 0.97 | <0.001 | 0.35 ± 0.03 | 0.36 ± 0.03 |
| | MD | 1, 44 | 0.018 | 0.08<sup>a</sup> | 0.102 | 7.18 ± 0.28 | 7.01 ± 0.23 |
| | Volume | 1, 44 | 0.237 | 0.39 | 0.029 | 10.03 ± 1.97 | 9.03 ± 2.28 |
| PoC-PrC<sub>hand</sub> u-fibers | LI FA | 1, 43 | 0.389 | 0.50 | 0.017 | 1.4 ± 6.9 | 2.8 ± 6.3 |
| | LI MD | 1, 43 | 0.747 | 0.90 | 0.002 | -0.2 ± 2.2 | 0.2 ± 2.5 |
| | LI Volume | 1, 43 | 0.125 | 0.29 | 0.051 | -2.0 ± 2.3 | 9.3 ± 30.1 |
| Left PoC-PrC<sub>hand</sub> | FC | 1, 38 | 0.127 | 0.29 | 0.06 | 1.30 ± 0.42 | 1.51 ± 0.50 |
| Right PoC-PrC<sub>hand</sub> | FC | 1, 38 | 0.045 | 0.13 | 0.102 | 1.12 ± 0.43 | 1.39 ± 0.46 |
| PoC-PrC<sub>hand</sub> | LI FC | 1, 38 | 0.336 | 0.47 | 0.024 | 16.2 ± 30.9 | 9.1 ± 27.7 |

Significant at <sup>a</sup> $q < .10$ .
Volume in ml, MD in ( $\times 10^{-4}$ mm<sup>2</sup>/s).

Using one-sample *t*-tests we observed significant leftward volume asymmetry of PrC_hand_ and PoC_hand_ in the TC (*p*=.011; Figure 1E) but not in the ASD group. For the laterality indices, there were significant diagnosis effects for both PrC_hand_ and PoC_hand_ volume, (ASD<TC; Figure 1E; Table 2). These group effects remained when additionally controlling for TBV, RMSD_rot_, and when we ran the analysis on only strongly right-handed individuals (EHI scores ≥80; remaining sample=17 ASD, 19 TC), to ensure the effect was not driven by weakly right-handed individuals (Supplementary Table 2).

Post hoc ANCOVAs on CT and SA were run to further investigate the regions that showed group differences in hand knob volume. There was a significant group effect in right PrC_hand_ SA (*p*=.012, partial η^2^=.129, ASD>TC), but not CT (*p*=.237, partial *η*^*2*^=.030). For the laterality indices these were not significant, however, small to medium group effects were observed in SA (PrC_hand_: *p*=.014, partial *η*^*2*^=.125; PoC_hand_: *p*=.063, partial *η*^*2*^=.073; both ASD<TC), in contrast to CT (PrC_hand_: *p*=.856, partial *η*^*2*^=.001; PoC_hand_: *p*=.238, partial *η*^*2*^=.030).

### Hand knob PoC-PrC structural and functional connectivity

We observed a significant diagnosis effect in MD of right PoC-PrC_hand_ tract, which was increased in the ASD compared to TC group (Figure 1F; Table 2). The effect remained when additionally controlling for tract volume (*p*=.017). Although MD for the left PoC-PrC_hand_ tract did not survive FDR correction, it was initially significant (p=.039) with a small effect size (Figure 1F; Table 2). This is noteworthy given that group differences in these u-fiber tracts bilaterally have been previously reported in ASD. We also observed a medium effect size of group in right PoC-PrC_hand_ FC, which was decreased in the ASD relative to TC group, but it did not survive FDR correction (Figure 1G; Table 2).

One-sample *t*-tests revealed leftward laterality of FA in the TC (*p*=.046; Figure 1E), but not in the ASD group, and leftward laterality of PoC-PrC_hand_ FC in the ASD group that was not found in the TC group. No group differences were found in LI of PoC-PrC_hand_ u-fiber or FC measures.

### Upper and lower pre and post-central volume and connectivity

The results of the upper and lower pre and postcentral measures are shown in Supplementary Table 2. No significant group differences survived FDR correction. While they did not reach the level of significance, a small effect size of group was observed in LI PoC_lower_ volume in the direction of greater leftward laterality in the ASD relative to TC group (partial *η*^*2*^=.071; Supplementary Figure 3). There were also medium effect sizes of group in MD of the right PoC-PrC_upper_ u-fiber tract (partial *η*^*2*^=.152, ASD>TC) and in FC of left PoC-PrC_lower_ (partial *η*^*2*^=.129; ASD<TC) and bilateral PoC-PrC_upper_ (left: partial *η*^*2*^=.145; right: partial *η*^*2*^=.126; both showing ASD<TC).

### Correlations between hand knob measures and motor skills

The results of the partial correlation analyses with the BMAT subscales are reported in Table 3. As BMAT Coordination scores were at ceiling for almost all TC participants, this measure was excluded from further analysis in the TC group. We observed several nominally significant associations in the ASD group, however, they did not survive FDR correction. Nonetheless these were medium effect sizes that have not been previously reported in the literature. These include positive correlations between LI PrC_hand_ volume and Manual Dexterity scores, and between right PoC_hand_ volume and Coordination scores, and a negative correlation between LI PoC-PrC_hand_ u-fiber FA and Coordination scores, in the ASD group (Table 3). No associations were found between PoC-PrC_hand_ FC and motor skills measures or with the Strength and Flexibility subscale in either group.

**Table 3.** Partial correlation results between hand knob measures and BMAT subscales in the ASD and TC groups.

|  | Manual dexterity |  |  | Coordination |  |  | Strength and flexibility |  |  |
| --- | --- | --- | --- | --- | --- | --- | --- | --- | --- |
|  | Partial *r* | *p* | *q* | Partial *r* | *p* | *q* | Partial *r* | *p* | *q* |
| ASD |  |  |  |  |  |  |  |  |  |
| Left PrC<sub>hand</sub> volume | 0.123 | 0.575 | 1.00 | -0.026 | 0.905 | 0.99 | 0.079 | 0.719 | 0.88 |
| Left PoC<sub>hand</sub> volume | 0.198 | 0.364 | 0.86 | 0.256 | 0.239 | 0.86 | 0.144 | 0.514 | 0.88 |
| Right PrC<sub>hand</sub> volume | -0.240 | 0.270 | 0.86 | -0.002 | 0.994 | 0.99 | 0.142 | 0.519 | 0.88 |
| Right PoC<sub>hand</sub> volume | 0.058 | 0.791 | 1.00 | 0.480 | 0.021<sup>a</sup> | 0.22 | 0.089 | 0.685 | 0.88 |
| LI PrC<sub>hand</sub> volume | 0.433 | 0.039<sup>a</sup> | 0.70 | 0.035 | 0.874 | 0.99 | -0.012 | 0.958 | 0.96 |
| LI PoC<sub>hand</sub> volume | 0.196 | 0.371 | 0.86 | -0.214 | 0.327 | 0.89 | 0.047 | 0.831 | 0.88 |
| Left PoC-PrC<sub>hand</sub> FA | -0.226 | 0.324 | 0.86 | -0.327 | 0.149 | 0.67 | -0.071 | 0.759 | 0.88 |
| Left PoC-PrC<sub>hand</sub> MD | -0.112 | 0.628 | 1.00 | 0.108 | 0.641 | 0.89 | 0.064 | 0.783 | 0.88 |
| Left PoC-PrC<sub>hand</sub> volume | 0.286 | 0.208 | 0.86 | 0.127 | 0.582 | 0.89 | 0.293 | 0.197 | 0.88 |
| Right PoC-PrC<sub>hand</sub> FA | -0.075 | 0.747 | 1.00 | 0.149 | 0.518 | 0.89 | -0.146 | 0.528 | 0.88 |
| Right PoC-PrC<sub>hand</sub> MD | -0.038 | 0.870 | 1.00 | 0.012 | 0.959 | 0.99 | 0.128 | 0.580 | 0.88 |
| Right PoC-PrC<sub>hand</sub> volume | 0.201 | 0.381 | 0.86 | 0.362 | 0.107 | 0.64 | 0.188 | 0.415 | 0.88 |
| LI PoC-PrC<sub>hand</sub> FA | -0.103 | 0.656 | 1.00 | -0.489 | 0.024<sup>a</sup> | 0.22 | 0.118 | 0.611 | 0.88 |
| LI PoC-PrC<sub>hand</sub> MD | -0.094 | 0.684 | 1.00 | 0.145 | 0.529 | 0.89 | -0.105 | 0.649 | 0.88 |
| LI PoC-PrC<sub>hand</sub> volume | 0.281 | 0.218 | 0.86 | -0.153 | 0.508 | 0.89 | 0.225 | 0.327 | 0.88 |
| Left PoC-PrC<sub>hand</sub> FC | 0.001 | 0.997 | 1.00 | 0.124 | 0.623 | 0.89 | -0.073 | 0.772 | 0.88 |
| Right PoC-PrC<sub>hand</sub> FC | -0.022 | 0.930 | 1.00 | 0.081 | 0.749 | 0.96 | -0.109 | 0.666 | 0.88 |
| LI PoC-PrC<sub>hand</sub> FC | 0.011 | 0.965 | 1.00 | 0.126 | 0.619 | 0.89 | 0.084 | 0.741 | 0.88 |
| TC |  |  |  |  |  |  |  |  |  |
| Left PrC<sub>hand</sub> volume | 0.088 | 0.688 | 0.83 | -- | -- | -- | 0.072 | 0.743 | 0.86 |
| Left PoC<sub>hand</sub> volume | 0.410 | 0.052 | 0.43 | -- | -- | -- | 0.127 | 0.565 | 0.86 |
| Right PrC<sub>hand</sub> volume | 0.395 | 0.062 | 0.43 | -- | -- | -- | 0.039 | 0.860 | 0.86 |
| Right PoC<sub>hand</sub> volume | 0.329 | 0.125 | 0.43 | -- | -- | -- | 0.170 | 0.439 | 0.86 |
| LI PrC<sub>hand</sub> volume | -0.298 | 0.167 | 0.43 | -- | -- | -- | -0.054 | 0.808 | 0.86 |
| LI PoC<sub>hand</sub> volume | -0.066 | 0.766 | 0.86 | -- | -- | -- | -0.094 | 0.670 | 0.86 |
| Left PoC-PrC<sub>hand</sub> FA | -0.226 | 0.572 | 0.79 | -- | -- | -- | -0.071 | 0.723 | 0.86 |
| Left PoC-PrC<sub>hand</sub> MD | -0.112 | 0.901 | 0.90 | -- | -- | -- | 0.064 | 0.298 | 0.86 |
| Left PoC-PrC<sub>hand</sub> volume | 0.286 | 0.453 | 0.79 | -- | -- | -- | 0.293 | 0.241 | 0.86 |
| Right PoC-PrC<sub>hand</sub> FA | -0.075 | 0.157 | 0.43 | -- | -- | -- | -0.146 | 0.171 | 0.86 |
| Right PoC-PrC<sub>hand</sub> MD | -0.038 | 0.883 | 0.90 | -- | -- | -- | 0.128 | 0.795 | 0.86 |
| Right PoC-PrC<sub>hand</sub> volume | 0.201 | 0.502 | 0.79 | -- | -- | -- | 0.188 | 0.682 | 0.86 |
| LI PoC-PrC<sub>hand</sub> FA | -0.103 | 0.142 | 0.43 | -- | -- | -- | 0.118 | 0.418 | 0.86 |
| LI PoC-PrC<sub>hand</sub> MD | -0.094 | 0.565 | 0.79 | -- | -- | -- | -0.105 | 0.237 | 0.86 |
| LI PoC-PrC<sub>hand</sub> volume | 0.281 | 0.636 | 0.82 | -- | -- | -- | 0.225 | 0.219 | 0.86 |
| Left PoC-PrC<sub>hand</sub> FC | -0.149 | 0.520 | 0.79 | -- | -- | -- | -0.046 | 0.844 | 0.86 |
| Right PoC-PrC<sub>hand</sub> FC | 0.172 | 0.456 | 0.79 | -- | -- | -- | -0.108 | 0.641 | 0.86 |
| LI PoC-PrC<sub>hand</sub> FC | -0.376 | 0.093 | 0.43 | -- | -- | -- | 0.068 | 0.770 | 0.86 |

<sup>a</sup> Findings with $p < .05$ uncorrected, and medium effect sizes.
<sup>b</sup> BMAT Coordination scores were at ceiling for almost all TC participants and was therefore excluded from analyses.

### Correlations between hand knob measures and autism symptoms

The results of the partial correlation analyses with the ADOS-2 sub-scores are reported in Table 4. After FDR correction, right PoC-PrC_hand_ u-fiber volume was negatively correlated with the Comparison score in the ASD group (Figure 2A). Associations with medium to large effect sizes were observed between right PoC-PrC_hand_ u-fiber volume and LI PoC-PrC_hand_ MD with the RRB sub-score (both showed a negative relationship), and between LI PoC-PrC_hand_ u-fiber FA and the Comparison score (positive relationship), but did not survive FDR correction (see Table 4).

**Table 4.** Partial correlation results between hand knob measures and ADOS-2 sub-scores in the ASD group.

|  | Social Affect |  |  | RRB |  |  | Comparison Score |  |  |
| --- | --- | --- | --- | --- | --- | --- | --- | --- | --- |
| | Partial $r$ | $p$ | $q$ | Partial $r$ | $p$ | $q$ | Partial $r$ | $p$ | $q$ |
| Left PrC<sub>hand</sub> volume | 0.191 | 0.382 | 0.74 | 0.022 | 0.921 | 0.92 | 0.076 | 0.730 | 0.88 |
| Left PoC<sub>hand</sub> volume | 0.087 | 0.692 | 0.78 | 0.083 | 0.705 | 0.92 | 0.046 | 0.835 | 0.88 |
| Right PrC<sub>hand</sub> volume | 0.201 | 0.357 | 0.74 | -0.197 | 0.367 | 0.66 | -0.066 | 0.766 | 0.88 |
| Right PoC<sub>hand</sub> volume | -0.151 | 0.492 | 0.74 | -0.165 | 0.452 | 0.74 | -0.308 | 0.152 | 0.46 |
| LI PrC<sub>hand</sub> volume | 0.115 | 0.601 | 0.77 | 0.336 | 0.117 | 0.42 | 0.263 | 0.225 | 0.46 |
| LI PoC<sub>hand</sub> volume | 0.249 | 0.251 | 0.74 | 0.229 | 0.294 | 0.66 | 0.295 | 0.172 | 0.46 |
| Left PoC-PrC<sub>hand</sub> FA | 0.224 | 0.330 | 0.74 | 0.035 | 0.880 | 0.92 | 0.243 | 0.289 | 0.52 |
| Left PoC-PrC<sub>hand</sub> MD | 0.024 | 0.917 | 0.94 | 0.040 | 0.863 | 0.92 | 0.084 | 0.718 | 0.88 |
| Left PoC-PrC<sub>hand</sub> volume | 0.198 | 0.390 | 0.74 | -0.214 | 0.352 | 0.66 | -0.028 | 0.903 | 0.90 |
| Right PoC-PrC<sub>hand</sub> FA | -0.143 | 0.537 | 0.74 | -0.309 | 0.173 | 0.52 | -0.279 | 0.221 | 0.46 |
| Right PoC-PrC<sub>hand</sub> MD | 0.158 | 0.494 | 0.74 | 0.356 | 0.113 | 0.42 | 0.345 | 0.125 | 0.46 |
| Right PoC-PrC<sub>hand</sub> volume | -0.268 | 0.240 | 0.74 | -0.505 | 0.019 | 0.17 | -0.584 | 0.005 | 0.09<sup>a</sup> |
| LI PoC-PrC<sub>hand</sub> FA | 0.289 | 0.204 | 0.74 | 0.425 | 0.055 | 0.33 | 0.493 | 0.023 | 0.21 |
| LI PoC-PrC<sub>hand</sub> MD | -0.108 | 0.641 | 0.77 | -0.540 | 0.012 | 0.17 | -0.372 | 0.097 | 0.46 |
| LI PoC-PrC<sub>hand</sub> volume | 0.222 | 0.335 | 0.74 | 0.223 | 0.332 | 0.66 | 0.274 | 0.229 | 0.46 |
| Left PoC-PrC<sub>hand</sub> FC | -0.019 | 0.939 | 0.94 | 0.052 | 0.837 | 0.92 | 0.064 | 0.802 | 0.88 |
| Right PoC-PrC<sub>hand</sub> FC | 0.199 | 0.428 | 0.74 | -0.049 | 0.848 | 0.92 | 0.119 | 0.637 | 0.88 |
| LI PoC-PrC<sub>hand</sub> FC | -0.239 | 0.339 | 0.74 | 0.149 | 0.556 | 0.83 | 0.058 | 0.818 | 0.88 |

<sup>a</sup> Significant at $q < .10$ .

**Figure 2.**
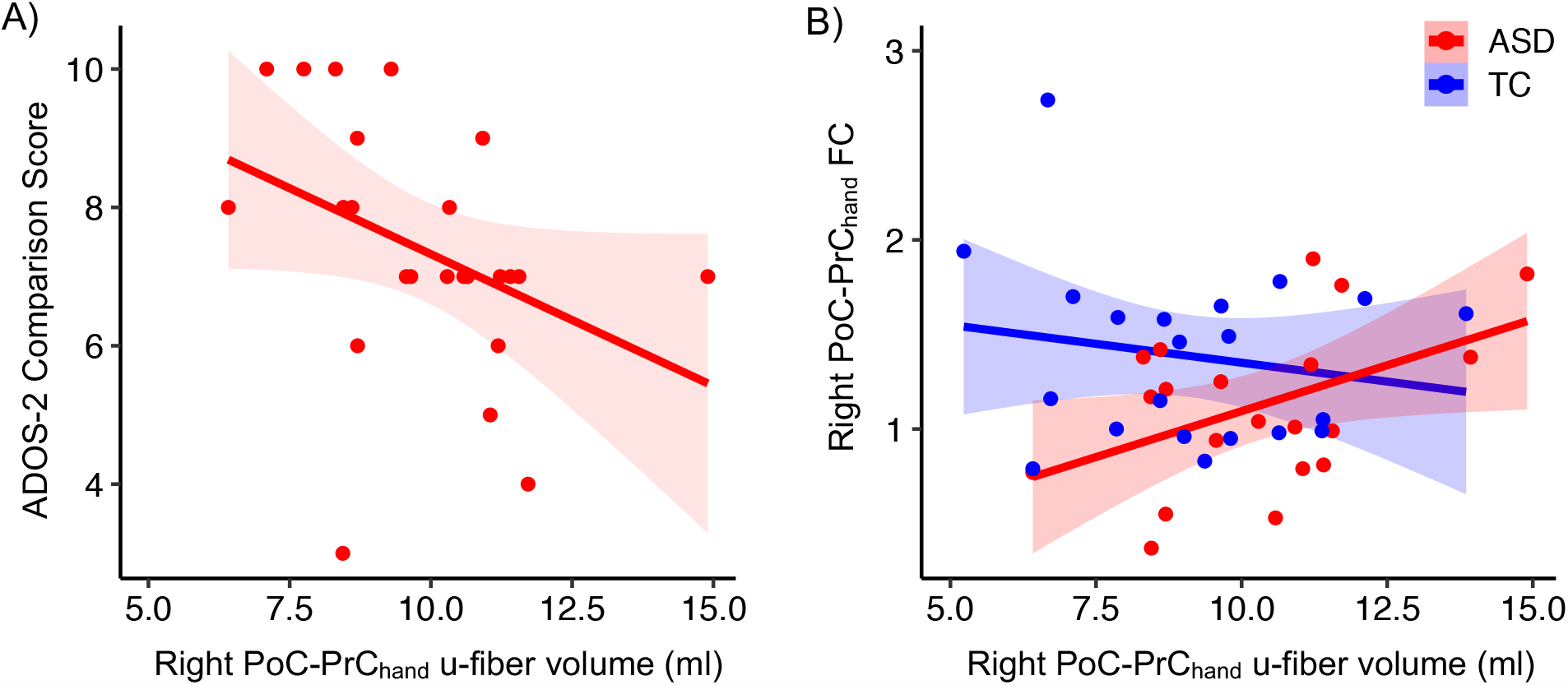
Scatterplots show A) a negative relationship between right PoC-PrC_hand_ u-fiber tract volume and current levels of autism severity as measured by the ADOS-2 Comparison Score (partial *r*=-.584, *q*=.09), and B) a positive relationship between right PoC-PrC_hand_ u-fiber tract volume and right PoC-PrC_hand_ FC, in the ASD group (partial *r*=.647, *q*=.06). Shaded areas in the scatterplots represent 95% confidence intervals.

### Correlations between PoC-PrC_hand_ structural and functional connectivity

After FDR correction, we found a significant positive association between right PoC-PrC_hand_ u-fiber volume and right PoC-PrC_hand_ FC in the ASD group only (partial *r*(16)=.647, *p*=.007, *q*=.06; Figure 2B). No associations between structural and functional connectivity measures were observed in the TC group.

## Discussion

To our knowledge, this is the first study to combine morphological, connectivity and asymmetry measures of the hand knob area in adults with ASD. Consistent with our hypotheses, we found that: 1) typical leftward asymmetry in PrC_hand_ and PoC_hand_ volume was reduced in adults with ASD, 2) for PrC_hand_, this reduced asymmetry was driven by increased right PrC_hand_ volume in the ASD group, and 3) PoC-PrC_hand_ structural connectivity was altered in ASD, with evidence of compromised microstructure (increased MD) in the right u-fibers. Together, our findings suggest a less lateralized neural circuitry as well as less support for efficient communication between sensory and motor regions of the hand knob area in middle-aged adults with ASD. While no associations between the hand knob measures and motor skills survived FDR correction, a negative relationship was observed between right PoC-PrC_hand_ u-fiber volume and autism symptom severity in the ASD group. This may indicate a compensatory role of the PoC-PrC_hand_ u-fibers in modulating motor behaviors associated with ASD.

### Reduced asymmetry of PrC_hand_ and PoC_hand_ in ASD

In typical individuals, handedness is associated with a deeper central sulcus, particularly in its dorsal portion, in the hemisphere contralateral to the dominant hand (Amunts et al., 1996, 2000; Cykowski et al., 2008; Klöppel et al., 2010; Sun et al., 2012). A deeper sulcus would presumably indicate an increased dorsal pre and postcentral cortical volume. Indeed, in this right-handed sample, we found that PrC_hand_ and PoC_hand_ volume were significantly left-lateralized in the TC group and this effect was primarily driven by SA rather than CT, consistent with previous findings. Higher rates of mixed or left-handedness reported in individuals with ASD, together estimated to be ∼42% when adjusting for publication bias (Markou et al., 2017) compared with ∼13% in the general population (Partida et al., 2020), would be expected to lead to reduced leftward or increased rightward volume laterality in the ASD population (i.e., consistent with our observed group effects). In our study, however, left-handed individuals were excluded and the diagnostic groups were matched on degree of handedness (i.e., EHI scores), to avoid skewing of group findings. As an added safeguard, we performed additional analyses restricted to the strongly right-handed individuals (excluding individuals with EHI<80). The group effects on hand knob volume and volume laterality, discussed below, had medium to large effect sizes in this more restricted sample, confirming that these results are not driven by the weakly right-handed individuals.

Our finding of increased right PrC_hand_ volume in middle-aged and older adults with ASD is consistent with previous reports of increased grey matter density in right pericentral cortex at the level of the hand knob in younger adults with ASD (DeRamus & Kana, 2015; Ecker et al., 2012). Our findings of reduced asymmetry in PrC_hand_ and PoC_hand_ volumes are also consistent with previous unilateral findings of increased right and/or decreased left grey matter density and volume in the pre- and postcentral gyri in ASD (Cauda et al., 2011; DeRamus & Kana, 2015; Ecker et al., 2012; Mahajan et al., 2016; Nickl-Jockschat et al., 2012; Säisänen et al., 2019), which may be interpreted in the context of reduced asymmetry. To our knowledge, morphological asymmetry has only been examined in a few studies on ASD (Floris, Lai, et al., 2016; Herbert et al., 2005; Postema et al., 2019). While none of these detected group differences in the precentral gyrus, Herbert et al. (2005) found reduced asymmetry in postcentral gyral volume in children with ASD.

As with language lateralization, functional lateralization of motor systems is widely viewed to be advantageous for neural communication by optimizing conduction time and wiring capacity (Karolis et al., 2019). Reduced asymmetry in morphology of the hand knob area suggests less hemispheric specialization for efficient neural motor processing of hand motor/premotor function in middle-aged adults with ASD. It may also reflect an altered strategy for motor processing over time, e.g., an increased reliance on specialized right hemisphere motor functions to correct and update ongoing actions (Mutha et al., 2012), in individuals with ASD. Indeed, increased recruitment of right pericentral and premotor areas during motor learning has been reported in young adults with ASD (Müller et al., 2004). Additionally, in healthy aging, a shift towards reduced functional hemispheric asymmetry including in the sensorimotor domain has been hypothesized and presumed to reflect compensatory changes (Cabeza, 2002).

By delineating the hand knob regions, we were able to perform analyses on different sub-parcels of the pre and postcentral gyri. We observed significant leftward volume asymmetry only in PrC_hand_ and PoC_hand,_ and not in the upper and lower sub-parcels in the TC group indicating that this asymmetry is largely specific to the hand knob region. This is consistent with previous morphometric studies in right-handed individuals (Amunts et al., 1996, 2000; Cykowski et al., 2008; S Rose et al., 2012). Thus, future studies investigating morphological asymmetry of the pre and postcentral gyri should take into consideration the likelihood of varying degrees (and possibly direction) of asymmetry along the dorso-ventral axis.

### Decreased PoC-PrC_hand_ u-fiber connectivity in ASD

Our findings also suggest altered microstructure of right PoC-PrC_hand_ u-fibers and a similar trend in left PoC-PrC_hand_ u-fibers. Increased MD in the ASD group, which may reflect axonal degeneration or possibly inflammation (Alexander et al., 2007), is in line with a previous report of increased MD (and reduced FA) of PoC-PrC_hand_ u-fibers bilaterally in younger adults with ASD (Thompson et al., 2017).

This potential reduction in structural connectivity between somatosensory (PoC_hand_) and motor (PrC_hand_) hand knob areas in ASD could have important functional implications. Diffusion tractography does not permit the distinction between afferent and efferent connections but bidirectional connections between primary somatosensory and primary motor cortex have been demonstrated using axonal tracing in mouse (Matyas et al., 2010), and these could have distinct functional relevance. Feedforward PoC_hand_-PrC_hand_ connections are likely to play an important role in learning new motor skills based on evidence in monkeys (Pavlides et al., 1993) and humans (Kumar et al., 2019; Veldman et al., 2018). Therefore, compromised microstructure of these u-fibers may contribute to abnormal patterns of motor learning in ASD (de Moraes et al., 2017). These u-fibers are likely also important for fine motor skills and impairment in these connections can affect grasp stability (Shinoura et al., 2005). Indeed, atypical grasp kinematics have been reported in adults with ASD (Cook et al., 2013). Future studies should directly test whether these u-fibers are linked to aspects of motor learning and fine motor control (e.g., handwriting difficulties (Grace et al., 2017)) in ASD, particularly in early childhood when development of these skills is most critical. Reciprocal PrC_hand_-PoC_hand_ connections may exert top-down influences on somatosensory cortex (e.g., M2 to S1 (Manita et al., 2015)) and could contribute to atypical sensory perception in ASD (Manita et al., 2015; Umeda et al., 2019).

One study found that FA of superficial white matter underlying the right pre and postcentral gyri positively correlated with finger tapping performance and that declining FA mediated age-related decline in fine motor performance in healthy adults aged 18-86 years (Nazeri et al., 2015). These u-fibers are thus particularly important for maintaining fine motor skills during aging.

### Correlations with motor skills

No significant correlations were found between hand knob measures and motor skills in either diagnostic group after correction for multiple comparisons. There were, however, correlations of medium effect size in the ASD group of hand knob measures with the Manual Dexterity and Coordination subscales, but not Strength and Flexibility. As expected, greater left-lateralized PrC_hand_ volume was linked to better manual dexterity skills, however, increased right PoC_hand_ volume and greater right-lateralized microstructural integrity (as measured by FA) of PoC-PrC_hand_ u-fibers tended to associate with better motor coordination skills in adults with ASD. Interestingly, Thompson and colleagues observed associations between manual dexterity skills (on a pegboard task) and right hemisphere u-fiber measures in their adult ASD sample, which contrasted with the left hemisphere u-fiber associations observed in their TC group (Thompson et al., 2017). Atypical right PoC_hand_ and u-fiber correlations with motor performance in adults with ASD could reflect an unusual reliance on on-line corrective sensorimotor processes in the right hemisphere.

### Correlations with autism symptom severity

Greater right PoC-PrC_hand_ u-fiber volume, reflecting more substantial structural connections, was significantly associated with lower severity of current autism symptoms (as measured by ADOS-2 Comparison Score) in middle-aged adults with ASD. While both RRB and Social Affect sub-scores showed a negative trend with right PoC-PrC_hand_ u-fiber volume, only the correlation with RRB was significant prior to FDR correction. One hypothesis is that the right u-fibers may be involved in individuals’ self-regulation of motor behaviors associated with ASD, such as repetitive movements (e.g., hand flapping, finger flicking). Furthermore, the observed positive correlation between right PoC-PrC_hand_ u-fiber volume and FC suggests that activity-dependent mechanisms are linked to changes in u-fiber tract volume and are behaviorally relevant in adults with ASD (Sampaio-Baptista & Johansen-Berg, 2017).

### Limitations

The small sample size of our study is one limiting factor. While our hand knob u-fiber findings replicates (and extends) a previous ASD study (Thompson et al., 2017), our novel laterality and morphology findings of the hand knob area in adults with ASD will require confirmation in larger samples. In particular, it is not known whether these findings are generalizable to left-handed adults with ASD. Lack of significant associations between the hand knob u-fiber measures and motor skills in the TC group may be due to a limited range of scores on the BMAT Short Form. Assessments which better capture the full range of motor function should be used in future studies. As intra-subject variability of motor task performance increases with aging (Hunter et al., 2016), repeat measurements may increase the reliability of motor measures in middle-aged and older adults.

## Conclusions

Our multi-modal MRI findings suggest that there is reduced morphological asymmetry and compromised PoC-PrC u-fibers of the hand knob area in middle-aged adults with ASD. Our findings may relate to atypical functional asymmetries found in children and young adults with ASD (Cardinale et al., 2013; Floris, Barber, et al., 2016), but could additionally reflect a compensatory functional overreliance on right hemisphere motor circuits over time. The right PoC-PrC_hand_ u-fibers were linked to their corresponding FC and may underlie compensatory self-regulation of unwanted motor behaviors seen in ASD. Given that manual motor function typically begins to decline in middle-age (Hoogendam et al., 2014; Ranganathan et al., 2001) and can significantly impact quality of life, longitudinal follow-up is needed.

## Data Availability

The data that support the findings of this study are openly available in the National Institute of Mental Health Data Archive (NDA) at https://nda.nih.gov/, reference number 2291.

## Acknowledgements

This study was supported by the National Institutes of Health R01 MH103494. The authors would like to thank all of the study participants.

## Supplementary Material

**Supplementary Table 1.** Frequency and proportion of hand knob shape variants by diagnostic group.

| Group | Shape | Left |  | Right |  | Overall |  |
| --- | --- | --- | --- | --- | --- | --- | --- |
|  |  | N | % | N | % | N | % |
| ASD | Omega | 20 | 80.0% | 20 | 80.0% | 40 | 80.0% |
|  | Epsilon | 4 | 16.0% | 5 | 20.0% | 9 | 18.0% |
|  | Null | 1 | 4.0% | 0 | 0.0% | 1 | 2.0% |
| TC | Omega | 20 | 83.3% | 19 | 79.2% | 39 | 81.3% |
|  | Epsilon | 4 | 16.7% | 4 | 16.7% | 8 | 16.7% |
|  | Null | 0 | 0.0% | 1 | 4.2% | 1 | 2.1% |

**Supplementary Table 2.** Results of ANCOVAs additionally controlling for TBV, RMSD_rot_ and excluding weakly right-handed individuals.

| Variable additionally controlling for | Region | Measure | df | $p$ | Partial $\eta^2$ |
| --- | --- | --- | --- | --- | --- |
| TBV | Right $\text{PrC}_{\text{hand}}$ | Volume | 1, 45 | 0.013 | 0.129 |
| | $\text{PrC}_{\text{hand}}$ | LI Volume | 1, 45 | 0.011 | 0.134 |
| | $\text{PoC}_{\text{hand}}$ | LI Volume | 1, 45 | 0.017 | 0.119 |
| $\text{RMSD}_{\text{rot}}$ | Right $\text{PrC}_{\text{hand}}$ | Volume | 1, 45 | 0.009 | 0.143 |
| | $\text{PrC}_{\text{hand}}$ | LI Volume | 1, 45 | 0.025 | 0.107 |
| | $\text{PoC}_{\text{hand}}$ | LI Volume | 1, 45 | 0.037 | 0.093 |
| Excluding weakly right-handed individuals | Right $\text{PrC}_{\text{hand}}$ | Volume | 1, 33 | <0.001 | 0.292 |
| | $\text{PrC}_{\text{hand}}$ | LI Volume | 1, 33 | 0.001 | 0.327 |
| | $\text{PoC}_{\text{hand}}$ | LI Volume | 1, 33 | 0.009 | 0.185 |

Abbreviations: $\text{RMSD}_{\text{rot}}$ =Root mean squared deviation in roll, pitch and yaw rotations from MNI standard space.

**Supplementary Table 3.**
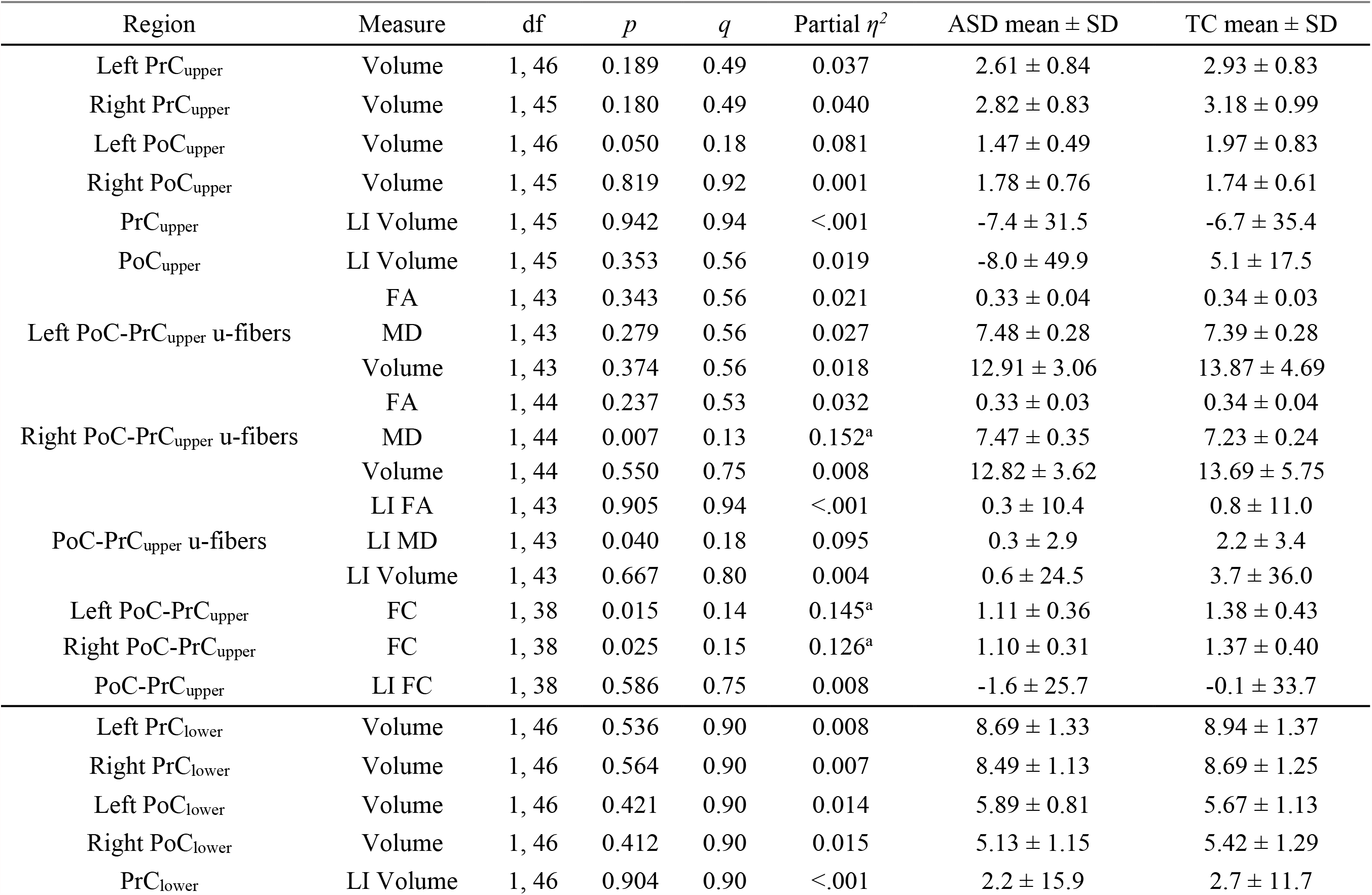
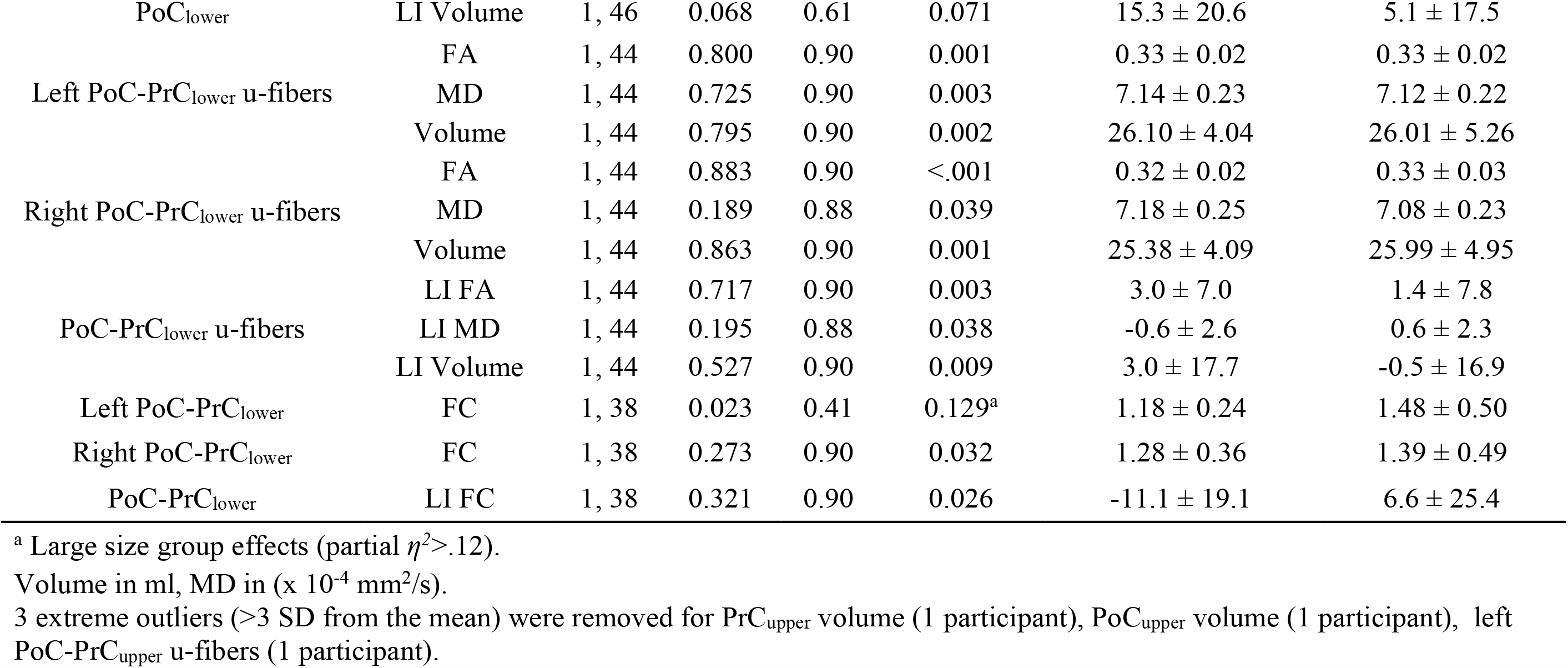
Results of the ANCOVAs performed on the upper and lower pre and postcentral measures and their group means, standard deviations.

**Supplementary Figure 1.**
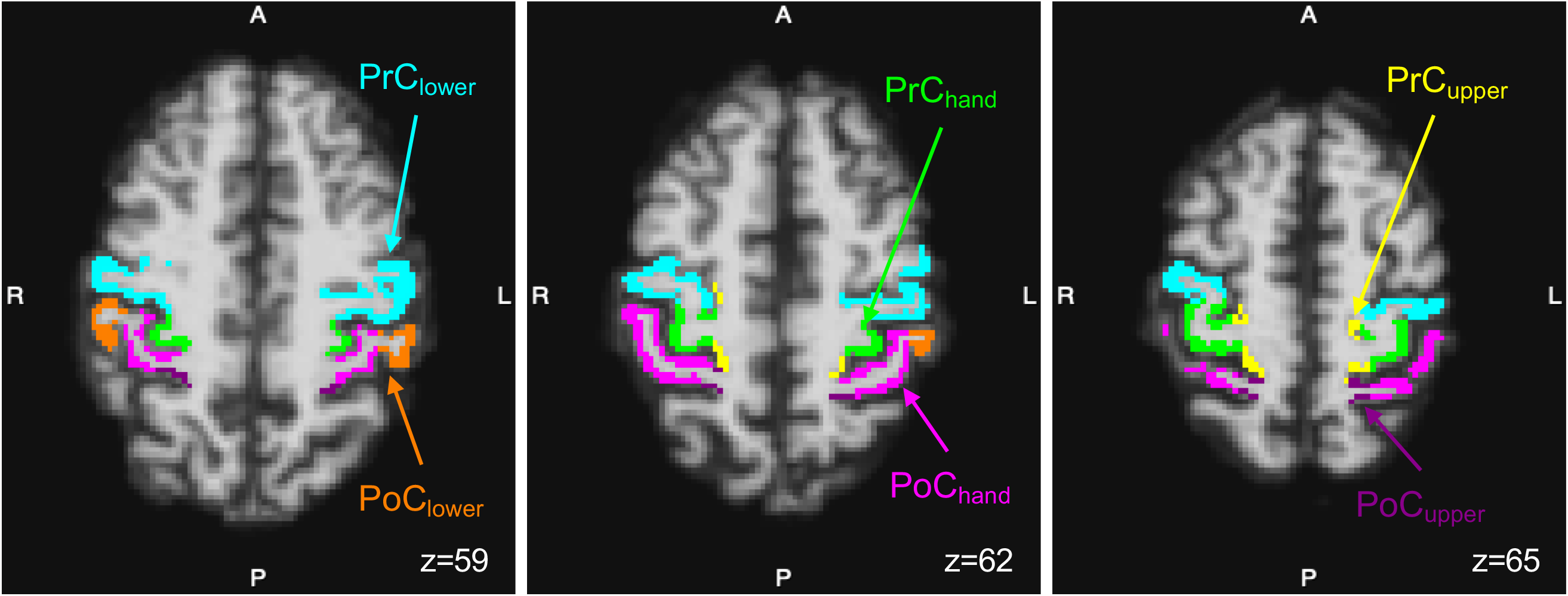
PrC and PoC sub-parcels shown on multiple axial slices in a representative participant with ASD.

**Supplementary Figure 2.**
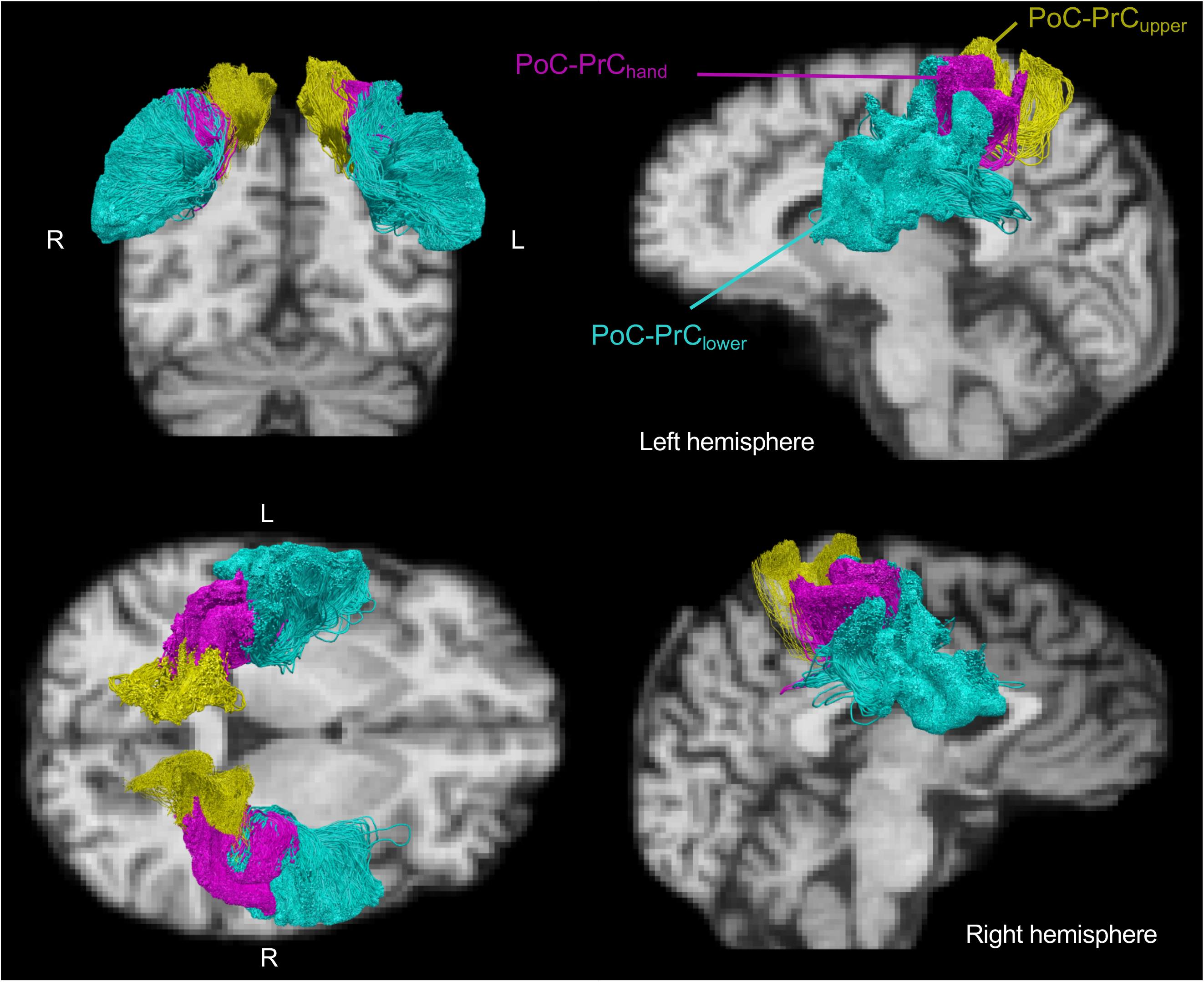
PoC-PrC u-fiber sub-tracts corresponding to PrC and PoC sub-parcels shown from multiple angles in native diffusion space with a T1-weighted underlay in a representative participant with ASD.

**Supplementary Figure 3.**
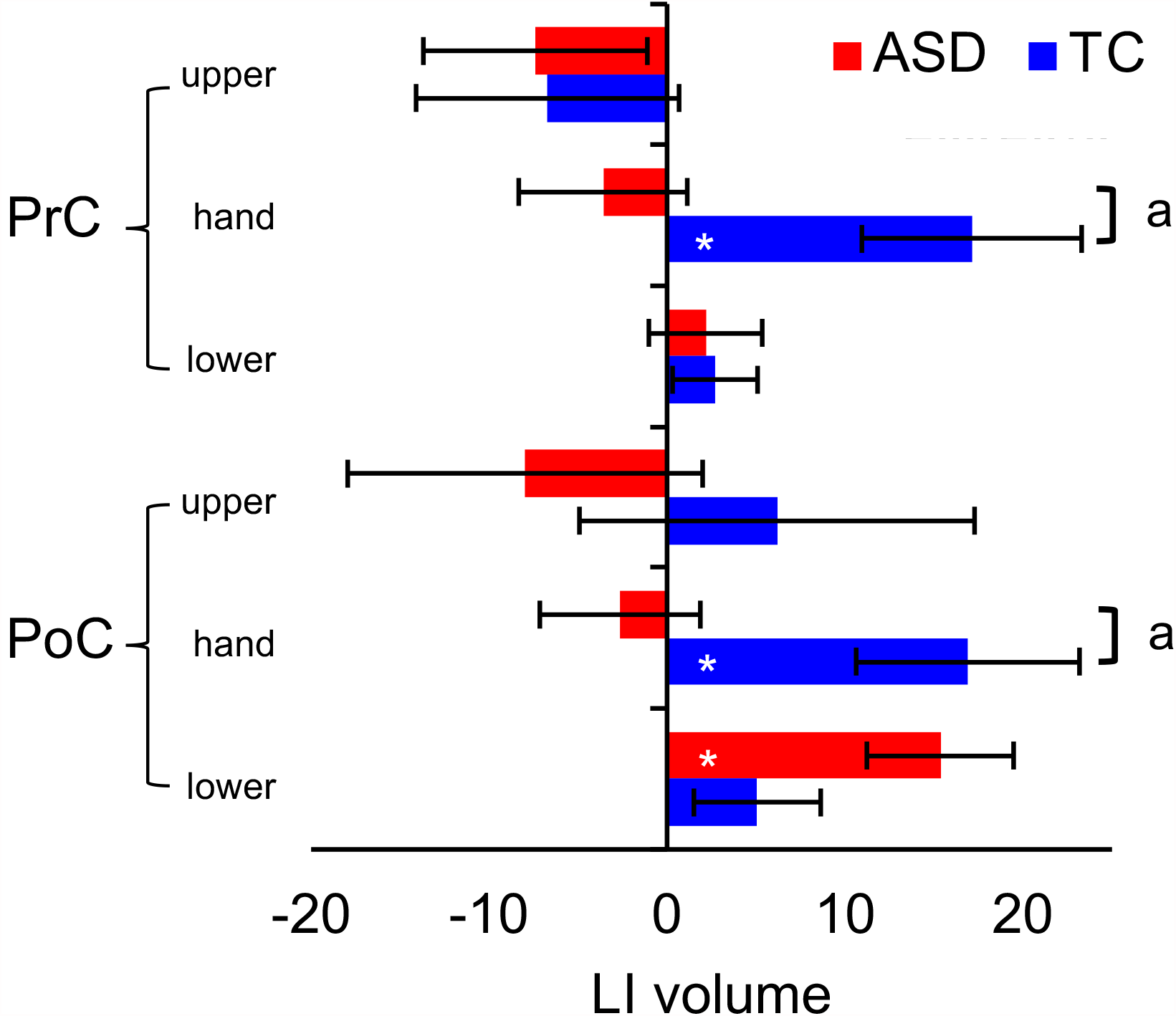
Laterality index of volume in the pre and postcentral sub-parcels (upper, hand knob, and lower). Significant at ^a^ *q*<.10 (group difference); ^*^*p*<.05, uncorrected (differs from zero).

